# Project ITCH: Cross-sectional surveys to understand residential vector control attitudes and practices across New England

**DOI:** 10.1101/2025.07.24.25332127

**Authors:** Fatma Tufa, Johanna Ravenhurst, Elissa S. Ballman, Neeta P. Connally, Janelle Couret, Nolan Fernandez, Allison M. Gardner, Jeff R. Garnas, William Landesman, Alexis L. White, Guang Xu, Stephen M. Rich, Andrew A. Lover, Thomas N. Mather

**Affiliations:** New England Center of Excellence in Vector-Borne Diseases, University of Massachusetts-Amherst, Amherst, MA, United States; Department of Biostatistics and Epidemiology, School of Public Health and Health Sciences, University of Massachusetts-Amherst, Amherst, MA, United States; School of Biology and Ecology, University of Maine, Orono, ME, United States; Department of Biology, Western Connecticut State University, Danbury, CT, United States; Department of Biological Sciences, University of Rhode Island, Kingston, RI, United States; Department of Natural Resources and the Environment, University of New Hampshire, Durham, NH, United States; Department of Natural Science, Vermont State University, Johnson, VT, United States; Department of Microbiology, College of Natural Sciences, University of Massachusetts – Amherst, Amherst, MA, United States; Department of Plant Sciences & Entomology, University of Rhode Island, Kingston, RI, United States

## Abstract

**Objective:** To understand the attitudes and prevalence of peridomestic vector control across New England, and to identify factors associated with self-reported residential vector control uptake.

**Methods:** An online-based survey captured 4,957 responses from households across New England in 2023 and 2024. Multivariable Poisson regression was used to quantify factors, including attitudes towards ticks and mosquitoes, motivations for vector control, and property characteristics, that are associated with total household residential vector control practices.

**Results:** The majority of respondents reported using at least one household-level vector control intervention in 2023 (88%) and 2024 (93%). Overall, higher levels of concern for ticks, mosquitoes, and mosquito-borne disease showed a gradient-response and were associated with statistically significant increasing rates of peridomestic vector control; however, concern for tick-borne disease was not associated with vector control usage (p-values > 0.05). After adjustment for socio-economics, yard size, property type, and levels of concern, Maine and Vermont both had lower incidence rates of total vector control interventions compared to the rest of New England [(aIRR = 0.89; 95% CI = 0.84 to 0.93; p-value = <0.001), (aIRR = 0.89, 95% CI = 0.82 to 0.96; p-value = 0.002), respectively]. Finally, protecting the health of family members had a strong association with peridomestic vector control uptake.

**Conclusions:** This study reveals factors associated with increased uptake of peridomestic vector control, including levels of concern, state of residence, and primary motivation for vector control use. Results support previous evidence and fill in gaps in geographic spread and the vector control interventions assessed. Future theory-based programming can help close gaps and increase uptake of peridomestic vector control. **Policy implications**: Differentials exist in state-level uptake of residential vector control interventions; mosquito-borne disease but not tick-borne disease, and protecting health of family members are associated with increasing rates of peridomestic vector control. Health promotion programs should incorporate these aspects in materials to foster appropriate levels of concern for all vector-borne diseases.

## I. Introduction

Vector-borne diseases (VBDs) are a major contributor to overall infectious disease burden in New England, and Lyme borreliosis is the most common VBD in the US, with an estimated 476,000 cases annually.^1^ The New England region reported 24% of the total national Lyme borreliosis cases in 2023, and 50% of all babesiosis cases in 2022. ^2,3^ The region is also endemic for Eastern Equine Encephalitis Virus (EEEV), which causes one of the most severe arboviral diseases in the US (13 cases in 2024; 77% of national total).^4^ Several other important vector-borne diseases are endemic in the region including anaplasmosis, Powassan virus and West Nile virus diseases, all of which have expanding geographic ranges.^5,6^

Prior studies have identified high-risk settings for VBD exposure, including peridomestic areas.^7,8^ Many interventions and products exist for homeowners to mitigate VBD risks in these settings, including removal of standing water, application of pesticides, deer fencing, and various vegetation modifications.^9–12^ However, there are very limited data on the current prevalence of these interventions, and little is known about the general attitudes of residential tick control across New England.

Prior studies have found inconsistent trends in the associations between reported levels of concern or geographic location and overall peridomestic vector control practices. Prior household surveys in MN, WI, MI, CT, and MD found no statistically significant associations between self-reported concern about Lyme disease specifically, and the use of pesticides on residential properties;^13,14^ whereas other studies in MD and TX found a significant association between concern about mosquito-borne disease (MBD) and the removal of standing water.^15,16^ In addition, a significant association between state/region (MI, MN, WI; NY; WI, NJ, NY; All regions in the US) and use of residential tick control methods were reported in three prior studies,^13,17–19^ while a third study found no differences among respondents from CT and MD.^14^

These prior studies generally had a more limited geographic focus, and were directed towards homeowners in counties with high Lyme disease incidence in a few specific states,^13,14^ or residing in a single locale,^15,17^ with the exception of two studies that evaluated all regions in the USA^19^ and compared a state in the Midwest (WI) with two states in the Northeast (NJ, NY).^18^ In addition, the self-reported residential vector control practices were limited in previous literature,^13–16^ with studies evaluating personal protective measures, a small number of peridomestic interventions,^17^ and either focusing solely on ticks or on mosquitoes. Consequently, there is a pressing need for research investigating factors that influence homeowner practices across the full range of residential vector control interventions relevant to both tick and mosquito control on a large geographic scale.

Deeper understanding of how geographic location and socio-economic factors impact homeowner practices could help to tailor future interventions to specific eco-epidemiological settings. Furthermore, several frameworks, including the Health Belief Model (HBM) and the Theory of Planned Behavior (TPB), provide theory-based methods for behavior change communication (BCC) programs.^20,21^ Previous studies have demonstrated impacts when using HBM as a framework to increase uptake of personal protection, illustrating successful application of these behavior change models to VBD programming.^22,23^ Furthermore, a study conducted in southeastern MA documented increased uptake of personal protection measures following participation in a theory-based educational program.^24^ In all, an *appropriate* level of concern about vector-borne disease risk is a potentially modifiable factor that might be targeted with future public health interventions to support increased uptake of residential vector control.

To address these gaps, the Centers for Disease Control and Prevention (CDC)-funded New England Center of Excellence in Vector-Borne Diseases (NEWVEC), implemented a region-wide attitudes and practices survey, Project ITCH (“Is Tick Control Helping?”). This study investigated potential associations between levels of concern about vector-borne diseases and geographic region on homeowner practices about residential vector control across New England. This is the first regional cross-sectional study to investigate factors influencing homeowner practices across a wide range of residential vector control interventions.

## II. Methods

### Survey instruments and study population

This cross-sectional study consisted of a 15-minute online survey implemented from March 2023 until October 2024 to assess attitudes and practices related to vector-control of New England residents. The survey instrument was developed by experts in tick and mosquito biology, ecology, and epidemiology, with investigators based in each of the six New England states: Maine, Vermont, New Hampshire, Massachusetts, Rhode Island, and Connecticut. Recruitment occurred through mass media sampling across all six participating states, including agricultural extension programs, local radio, newsletters, social media, and other media outlets. The primary investigator in each state led local recruitment efforts, which allowed for setting-specific approaches. Survey promotion was concentrated from March to early May, prior to peak nymphal *Ixodes scapularis* (blacklegged tick) activity in New England; survey participants had to be 18 years of age or older. Responses were collected using the REDCap platform, hosted at the University of Massachusetts Chan Medical School.^25^

The survey instrument spanned six domains: i. general outdoor activities and household exposure to vector-borne diseases; ii. housing type; iii. general attitudes/perceived risk; iv. personal protection practices and motivation; v. current residential vector control practices and attitudes; and vi. household demographics. Queries included a combination of check-all-that-apply, Likert scales, multiple choice, and free-form responses. In April 2024 the survey was revised to include a new screening questionnaire for a follow-up tick-surveillance study, and edited for clarity based on feedback from participants and research staff. Secondly, residential vector control options were separated into queries for interventions related to ticks and to mosquitos (which were previously combined).

The study protocol was reviewed and approved by the University of Massachusetts Amherst Institutional Review Board (IRB) as the central IRB (Protocol # 3969). Approved materials were shared with all partner institution IRB offices, with any additional approvals as required by each partner institution. Participants provided electronic informed consent via REDCap.^25^

### Data cleaning and quality checks

Data quality reports were reviewed throughout implementation to assess any issues with design or clarity of survey questions. Prior to analysis, records not meeting inclusion criteria were removed. The primary outcome for this analysis was the self-reported residential vector control practices, from a check-all-that-apply query. For analysis, the uptake of residential control actions was summarized in two complementary ways: as a binary outcome (any vector control/no vector control), and then as the sum of all self-reported vector control interventions, including both ticks and mosquitoes. The category of “any other” vector control interventions was excluded in the analysis, due to low response (1.2% for ticks and 1.6% for mosquitoes in 2024; 2023 did not have an “any other” option).

### Statistical analysis

Exploratory data analyses were performed for total survey, and by individual state. Associations between categorical covariates and use of any vector control intervention (as a binary) or state were assessed using χ^2^ or Fisher’s exact tests; continuous variables were assessed using the non-parametric Wilcoxon rank-sum test due to departures from normality.

Two multivariable models were developed to quantify factors associated with vector control interventions. Robust Poisson models^26^ for binary outcomes were used to estimate prevalence ratios. The main outcome (dichotomous use of “any vector control” intervention) was very common in the survey, potentially leading to biased estimates in a standard logistic regression, and justifying the use of a Robust Poisson model. In parallel, a Poisson model for discrete counts was used for the total number of self-reported interventions; this model assumes exchangeability (equivalence) between all potential interventions in the survey. Negative binomial regressions showed no improvement over Poisson models. All exploratory variables with prior reporting in the literature and/or biological plausibility, and with p-values ≤ 0.2 were included in initial models.^27^ LASSO (least absolute shrinkage and selection operator) was then used to construct final candidate models,^28^ with adjustment for gender, income, state of residence, yard size, property type and respondent age. Collinearity was assessed; where detected, additional checks using AIC were performed, with the lowest AIC model being retained. Several Likert-scale items had very low responses for “Strongly disagree” (< 5%), which led to instability from quasi-separation. Penalized likelihood estimation (brglm2 in R ^29^) was used to address this; two lowest levels were also combined (without change by AIC). Finally, Nagelkerke’s R^2^ and standardized residual plots were examined to assess model fit.^30^ Data were analyzed with SAS 9.4 (SAS Inc., Cary, NC, USA); or R version 4.4.2 (*Pile of Leaves*); all tests were two-tailed, with ɑ = 0.05.

## III. Results

### Demographics of respondents

From March 2023 to October 2024, a total of 8,595 responses were recorded. Of these, 1,865 (22%) were excluded due to an incomplete informed consent; 1,113 (13%) were excluded due to missing responses; 221 (3%) were duplicate responses; and 439 (5%) were excluded due to living outside of New England, not owning their home, or being unable to make property-related decisions. The final analytical sample includes responses from 4,957 households; demographics of which shown in Table 1. The survey had broad coverage across all regions of New England, with the exception of very sparsely populated areas in Northern ME and NH (Fig 1<u>, Panel A</u>). The study sample was predominantly female (69%), white (93%), and generally had high educational attainment, with 49% reporting a graduate degree. The reported household median income (> $100,000 - 200,000 category), was higher than the New England household median ($92,017).^31^

**Fig 1.**
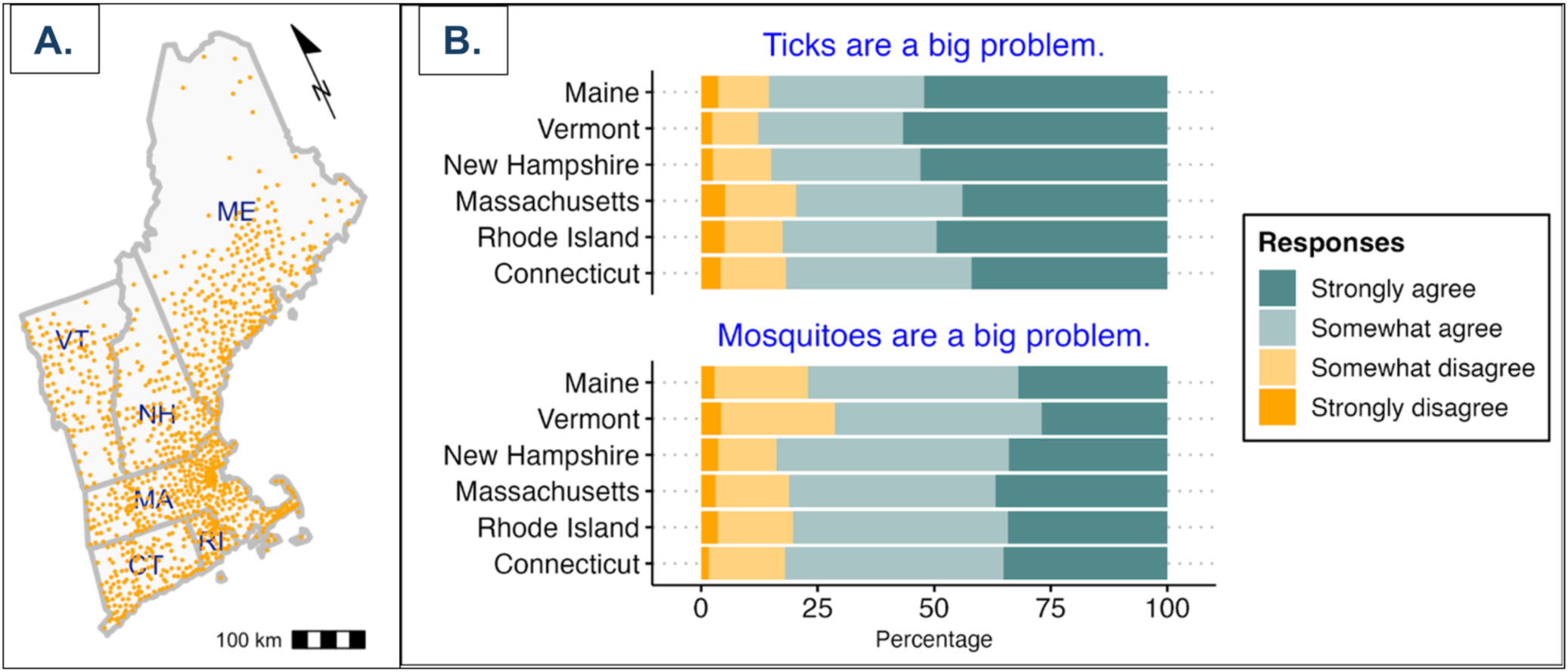
(**Panel A**). Reported household locations, by postal codes, “Is Tick Control Helping” (ITCH) survey respondents, New England, USA, 2023-2024. (N = 1,130 postal codes). (**Panel B**). Response to Likert-scale questions about vectors in domestic settings,“Is Tick Control Helping” (ITCH) survey respondents, New England, USA, 2023-2024 (N = 4,919).

**Table 1.**
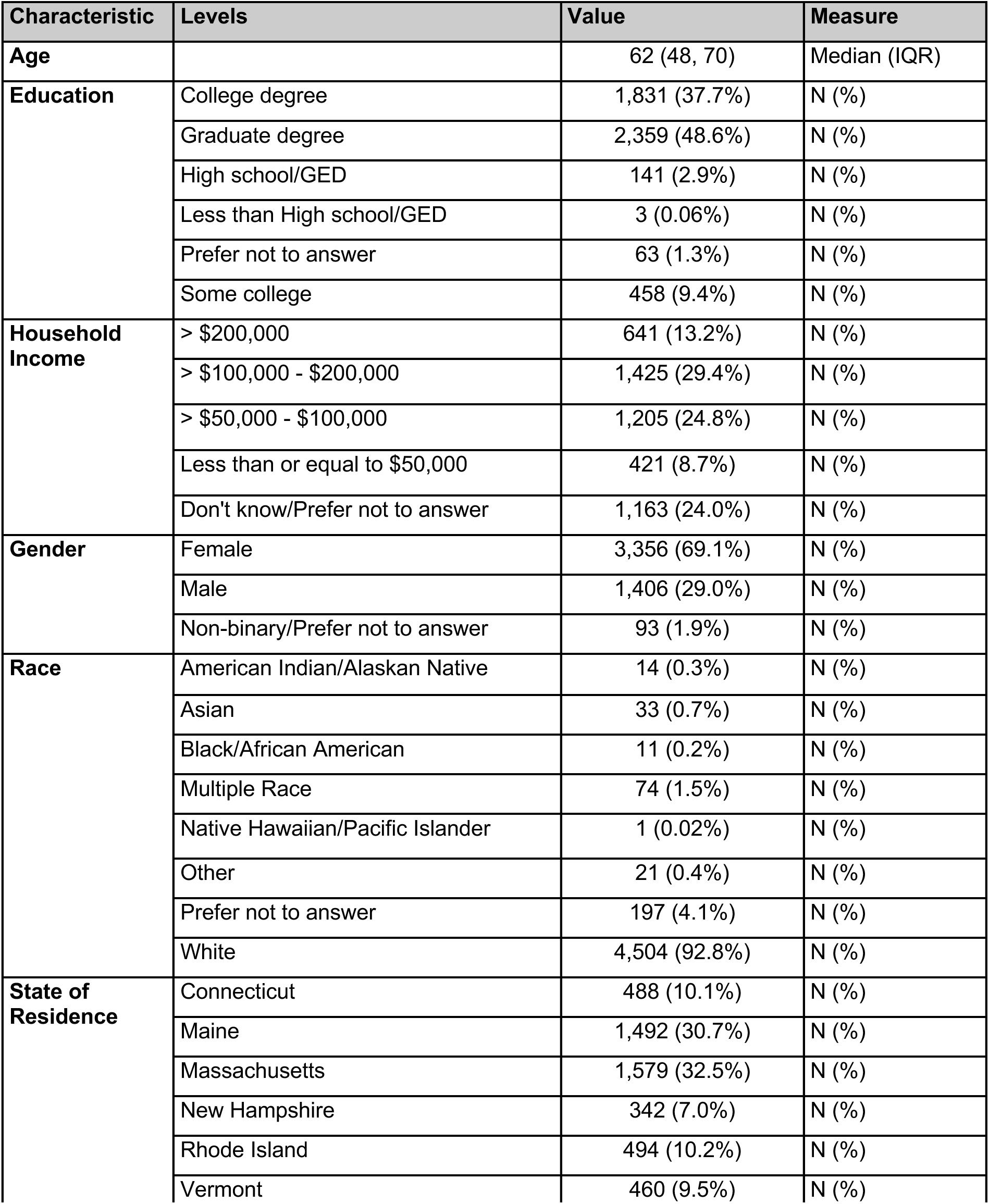
Overview of “Is Tick Control Helping” (ITCH) survey respondents, New England, USA, 2023-2024 (N = 4855).

#### Self-reported prevalence of residential vector control

Across the survey, the majority of households reported using at least one type of vector control intervention in both 2023 (88%) and 2024 (93%). For specific interventions, there was wide variation in prevalence, but generally limited variability across states. For several interventions there was very high usage; most households reported removing items that store water around the property in 2023 (ME: 53.7%, NH: 70.3%, VT: 61.9%, RI: 69.3%, MA: 74.3%, CT: 70.2%) and 2024 (ME: 70.0%, NH: 69.0%, VT: 37.5%, RI: 69.2%, MA: 74.5%, CT: 62.3%), with a drop in uptake in VT in 2024. A large number of households also reported vegetation and leaf removal. Several interventions were very infrequent across all states, with deer fencing, tick tubes, and rodent bait boxes reportedly having ≤ 20% uptake in both 2023 and 2024 (<u>See Supplemental Information, Tables S1A and S1B</u>).

Overall use of peridomestic vector control varied widely across states; as commercial pesticide application, where in 2023 Vermont had a lower prevalence (2.7%) compared to other states (ME: 12.3%, NH: 13.1%, RI: 31.3%, MA: 16.2%, CT: 20.2%). In 2023, all states had a median of 2.0 interventions, with the exception of Connecticut (3.0). Using the expanded 2024 survey questions, a wider range of values was recorded (medians from 1.5 to 3.0; Table 2). Importantly, while there was limited variability in median values, several states had sub-populations with very high uptake of vector control options (<u>See Supplemental Information, Figure S1</u>).

**Table 2.** Prevalence and total number of self-reported household-level vector control actions, New England, 2023-2024. (N =4,855).

| State | 2023 survey |  |  |  | 2024 survey |  |  |  |
| --- | --- | --- | --- | --- | --- | --- | --- | --- |
|  | Prevalence any vector control actions |  | Total reported vector control actions |  | Prevalence any vector control actions |  | Total reported vector control actions |  |
|  | n | (%) | Median | IQR | n | (%) | Median | IQR |
| Connecticut | 178 | (93.8) | 3.0 | 2.0 | 310 | (89.4) | 2.0 | 2.0 |
| Maine | 1,482 | (83.1) | 2.0 | 2.0 | 10 | (90.0) | 2.0 | 2.0 |
| Massachusetts | 1,140 | (91.8) | 2.0 | 2.0 | 439 | (94.3) | 2.0 | 2.0 |
| New Hampshire | 313 | (90.1) | 2.0 | 2.0 | 29 | (100.0) | 3.0 | 3.0 |
| Rhode Island | 361 | (90.9) | 2.0 | 2.0 | 133 | (97.7) | 2.0 | 2.0 |
| Vermont | 452 | (85.0) | 2.0 | 2.0 | 8 | (75.0) | 1.5 | 2.0 |

#### Self-reported prevalence of personal protective measures

Overall, for most all personal protection measures, the majority of households self-reported they “sometimes” used it during outdoor activities. For a few of these, including tick checks and treating pets, the majority of households reported “always” performing the action. The full distributions of self-reported personal protective measures can be found in the supplemental information (Tables S2A and S2B).

### Measures of perceived risk (attitudes)

Five-point Likert scales were used to assess perceived levels of risk for both MBD and TBD. Across all responses, many participants “strongly agreed” that ticks are a big problem around their house (49%) and also “Strongly agreed” about being concerned about TBD (78%). Levels of concern were generally lower for MBD compared to TBD, with 45% of participants reporting they “Strongly agreed” that mosquitoes were a big problem around their house, and 46% reported “Strongly agreed” to being concerned about MBD. Across states in New England, there is strong evidence for differences in concern about ticks (Fisher’s exact test, p-value < 0.0001) and mosquitoes (p-value < 0.001). Higher levels of concern are seen in the Northern regions within New England, from respondents who “Strongly agreed” ticks are a problem (Vermont, 56%; New Hampshire, 52%; and Maine 52% vs. Connecticut, 42%; Massachusetts 44%), with Rhode Island leaning towards the middle (49%), see more in Fig. 1<u>, Panel B</u>.

### Changes in behavior (practice)

There were statistically significant differences in responses to changing outdoor activities due to ticks and mosquitoes. More households “somewhat agreed” about changing their outdoor activities in response to ticks (31%) and mosquitoes (40%) compared to those that “Strongly agreed” to changing their outdoor activities due to ticks (25%) and mosquitoes (20%). Significant differences were also found between levels of concern for ticks and changing outdoor activities due to ticks (χ^2^ test, p-value <0.001). For example, 70% of respondents “Strongly agreed” to changing their outdoor activities due to ticks and that ticks are a problem around their house, whereas 2.2% “Strongly agreed” to changing their outdoor activities due to ticks and “Strongly disagreed” ticks are a problem around their house. Similarly, significant differences were found between levels of concern for mosquitoes and changing outdoor activities due to mosquitoes (χ^2^ test, p-value <0.001).

### Self-reported vector-borne disease in households

TBD incidence was unexpectedly common across the entire sample, with 44% of all households self-reporting having a household member ever been diagnosed with a TBD. Stratifying by time periods, 12% of all respondents reported having a diagnosis within the past year; 25% were diagnosed from 1-5 years ago; and 29% had a diagnosis more than 5 years ago. In contrast, the rates of self-reported MBD were very low across the survey, with only 1% of participants reporting a household member ever having been diagnosed with any MBD.

### Factors associated with household use of vector-control interventions

Two complementary multivariable models were used to quantify factors associated with household-level vector control usage. After adjusting for demographics, yard size, and year of survey, the factors significantly associated with use of total vector control interventions included: the household’s primary motivation for vector control activities; self-reported higher levels of concern for ticks, mosquitoes, and for mosquito borne disease; any previous diagnosis of a TBD; and state of residence. Here, we focus on the Poisson count model for total vector control interventions, where unadjusted and adjusted analyses are shown in Table 3; the robust Poisson model for unadjusted and adjusted prevalence ratios showed largely consistent results, but with limited discrimination (<u>See Supplemental Information, Table S3)</u>.

**Table 3.**
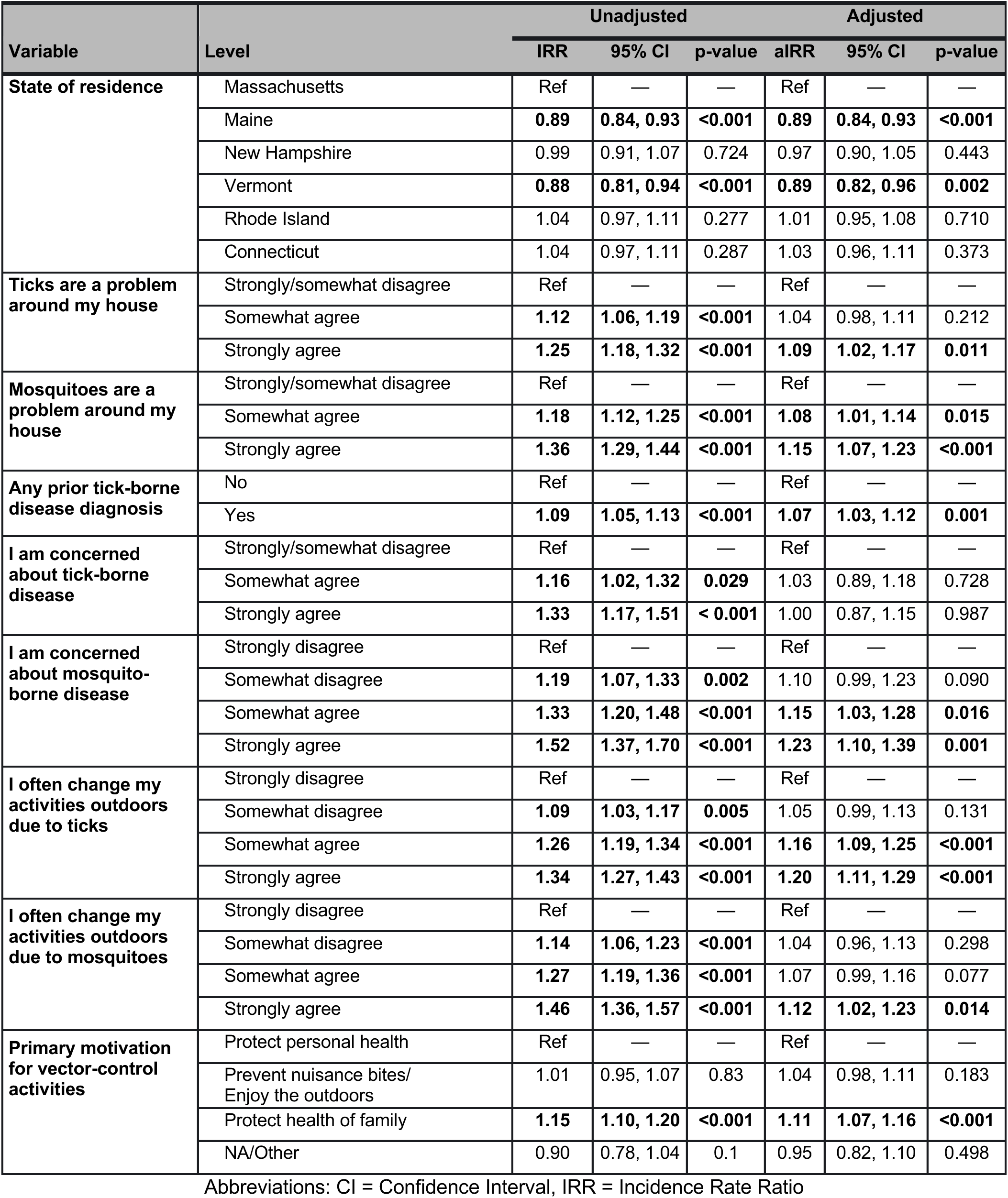
Multivariable Poisson models for total self-reported vector control interventions at the household level, New England, 2023-2024. Notes: IRR = incidence rate ratio; final model adjusted for age, survey year, sex/gender, yard size, and household income. Values in boldface are statistically significant with p < 0.05.

Households that “Strongly agreed” or ”Somewhat agreed” to changing their activities outdoor due to ticks had 20% and 16% higher incidence rates of total vector control interventions adjusted for covariates [(aIRR = 1.20, CI = 1.11, 1.29, p < 0.001), (aIRR = 1.16, CI = 1.09, 1.25, p-value < 0.001), respectively] relative to those that “Strongly disagreed”.

Survey respondents who “Strongly agreed” that ticks were a problem around their house had a 9% greater incidence rate of total vector control interventions adjusted for covariates (aIRR = 1.09, CI = 1.02, 1.17, p-value = 0.0011) relative to those that “Strongly disagreed”. In contrast, higher levels of concern about tick-borne disease (as opposed to ticks specifically) were not associated with increased uptake of vector-control interventions.

Households that “Strongly agreed” about changing their outdoor activities due to mosquitoes had 12% higher incidence rate of total vector control interventions (aIRR = 1.12, CI = 1.02, 1.23, p-value = 0.014) than those who “Strongly disagreed”, adjusted for covariates. Those that “Strongly agreed” or ”Somewhat agreed” to being concerned about MBD [(aIRR = 1.23, CI = 1.10, 1.39, p-value = 0.001) and (aIRR = 1.15, CI = 1.03, 1.28, p-value = 0.016, respectively]; and “Strongly agreed” or “Somewhat agreed” to mosquitoes being a problem around their house had 15% and 8% higher incidence rate (aIRR = 1.15, CI = 1.07, 1.23, p-value < 0.001 and aIRR = 1.08, CI = 1.01, 1.14, p-value = 0.015, respectively) of total vector control interventions than those who “Strongly disagreed” or “Somewhat disagreed”, adjusted for covariates.

Households whose primary motivation for vector-control activities was “protecting the health of their family” had a 11% higher incidence rate of total vector control interventions (aIRR = 1.11, CI = 1.07, 1.16; p-value < 0.001) relative to survey respondents whose primary motivation was “protecting personal health”, after adjustments for baseline demographics.

Households who responded “Yes” to having a prior TBD diagnosis had 7% higher incidence rate of total vector control interventions (aIRR = 1.07, CI = 1.03, 1.12, p-value = 0.001) than households that responded “No” to having any prior TBD diagnosis in their household, adjusted for covariates. Additionally, with Massachusetts as the reference, residents of Maine and Vermont had 11% lower incidence rate of total vector control interventions [(aIRR= 0.89; CI = 0.84, 0.93, p-value < 0.001) and (aIRR= 0.89; CI = 0.82, 0.96, p-value = 0.002), respectively), while there was no evidence for differences for the other four states (all p-values > 0.05).

To assess differences between the three northern states versus the three southern states, an additional model with these categories was explored. In this model, after adjustment for a range of factors (including income, education, yard size, property type, and the levels of concern) with the three Southern states (MA/CT/RI) as reference, residents of the three northern states had 10% lower incidence of vector control incidence rate of total vector control interventions (aIRR = 0.90; CI = 0.86, 0.94, p-value < 0.001). (<u>See Supplemental Information, Table S4</u>).

## IV. Discussion

Results from this study bridges gaps in understanding of attitudes and residential vector control practices across New England. Overall, households revealed high levels of concern for ticks, mosquitoes, and MBD, with a large number of respondents changing their activities outdoors due to ticks and mosquitoes. Additionally, the self-reported primary motivation for vector control programming for households was protecting the health of the family with almost half of respondents reporting a tick-borne disease diagnosis from a household member at some point in their life.

The majority of New England residents surveyed in this study reported using some type of residential vector control in 2023 and 2024. A large proportion of residential vector control interventions included removing standing water (to control mosquitoes) and vegetation removal (to control ticks and/or mosquitoes). For personal protective measures, households self-reported “sometimes” performing most personal protection measures to reduce their general risk of vector-borne disease, while tick checks and treating pets with acaricides were “always” performed. Such personal protective measures were also seen in other studies, where tick checks and treating pets were commonly reported.^14,32,33^ Further analysis of these items will be reported in future work.

In fully-adjusted models, relative to Massachusetts, two states (ME and VT) have statistically significantly lower uptake of residential vector control interventions. The range of covariates adjusted for include income, yard size, TBD/MBD concern, and recent TBD diagnosis. This finding implies, that in general, at the same levels of socio-demographics, survey respondents in ME and VT are significantly less likely to use vector control interventions in peridomestic settings. The reasons for this might include more limited access to vector control interventions (due to recent expansion of vector ranges), or feasibility in very different ecologies (not captured in income/socioeconomic status/yard size). This suggests that vector control programming is not “one size fits all” and will need to be tailored to the specific socio-demographic and ecological settings across New England.

Adjusted models also found that households that generally had high levels of concern for ticks, mosquitoes, and MBD had a greater incidence rate of using more total vector control measures. This finding is consistent with two other studies, where high levels of concern about MBD increased the likelihood of removing standing water,^15,16^ and likely due to rapidly changing risks associated with both ticks and with mosquitoes. Many areas in Northern New England have reported increasing caseloads from TBD and an increasing geographic range of ticks.^34^ Additionally, the recent emergence of EEEV in Northern New England, shifting the needs for vector control programming in the region, potentially due to changing avian migration patterns,^35^ has required expansion of regional vector surveillance programs.

Overall, the strongest association with vector control uptake in this study is self-reported concern about mosquito-borne disease, which shows a clear gradient with increasing rates of vector control usage. This exposure-response strengthens the validity of the survey items in capturing variability relationships between levels of concern and uptake of interventions as per Hill’s criteria (which may or may not be causal). There are several potential explanations for the contrasting lack of association between vector control uptake and levels of TBD concern. The most publicized mosquito-borne diseases (WNV disease and EEE) generally cause severe disease and/or mortality, whereas Lyme borreliosis, generally causes more limited illness. In addition, the differences in cost and accessibility of tick and mosquito control can lead to this lack of association (e.g., commercial pesticide application vs. removing standing water). This discordant response suggests an important gap in population-level risk assessments (prioritizing responses to disease severity vs infection frequency). Health messaging and behavior change communication should address this gap, and work to develop appropriate levels of concern for all types of vector-borne infections.

Finally, the finding that the priority motivator for taking action is to protect the health of family members represents a key insight for well-targeted BCC messaging.^36^ While studies evaluating BCC for TBD and for domestic MBD are limited, extensive programmatic data exists for other VBD, including malaria. A recent systematic review of malaria BCC programs concluded that programs that integrate mass education, interpersonal education, and the involvement of specialists in health instructional strategies is critical for maximizing impact.^37^ Applying evidence-based health behavior theories including HBM and TPB, while incorporating the prioritization of protecting the health of families, can aid in the implementation of residential vector control programming, help address regional gaps, and increase uptake of interventions to reduce ticks, mosquitoes, and associated infections.

The study population largely consisted of well-educated, higher income, and female participants, with high levels of vector control usage. Additionally, a large proportion of households had someone that had ever been diagnosed with a TBD. This suggests that the respondents to the survey are at highest-risk for TBD, and leaves a gap for information on households who implement fewer residential vector control actions.

### Strengths and limitations

A major strength is the large sample size with wide coverage across the Northeast, supported by broad mass media exposure. Households were asked about a variety of vector control interventions on their property, rather than only focusing on pesticide applications or standing water removal as in previous studies.^13–16^ While not a fully-validated instrument, this survey was developed by vector-borne disease experts from across the region, ensuring good alignment with regional trends.

This study has several limitations. The mass media sampling strategy varied by state, with the largest proportion of responses being in Massachusetts and Maine. In addition, people with high levels of concern about vector-borne diseases may be more likely to participate, introducing possible selection bias. More importantly, the outcome of the total number of vector control interventions implies that all vector control interventions are exchangeable (having equivalent impact), which is an oversimplification as there are large differences in both the effectiveness and in the costs associated with different interventions for ticks, and mosquitoes, and VBD.^9,12^

## Conclusions

This study identified factors associated with increased uptake of residential vector control interventions, as well as measured regional variation in attitudes on ticks, mosquitoes, and MBD and its associated residential vector control uptake. Multivariable regression models support the influence of levels of concern on residential vector control, supporting previous evidence, and fill gaps in the literature regarding geographic coverage and the variety of vector control interventions evaluated. Results suggest the application of theory-based educational vector control programming can help increase routine and regular uptake of residential vector control practices in peridomestic settings, tailored to geographic location and its associated eco-epidemiological settings.

## Ethics statement

IRB approval was obtained (UMass reference number 3969; Jan. 27, 2023), with reliance agreements to partner sites as required.

## Funding statement

This research was generated from the NEWVEC/UMass Amherst, which is funded by the cooperative agreement U01CK000661 from the Centers for Disease Control and Prevention. The contents are solely the responsibility of the authors and do not necessarily represent the official views of the Centers for Disease Control and Prevention.

## Data Availability

All data produced in the present study are available upon reasonable request to the authors.

## Supplemental Information/Appendix

**Fig S1.**
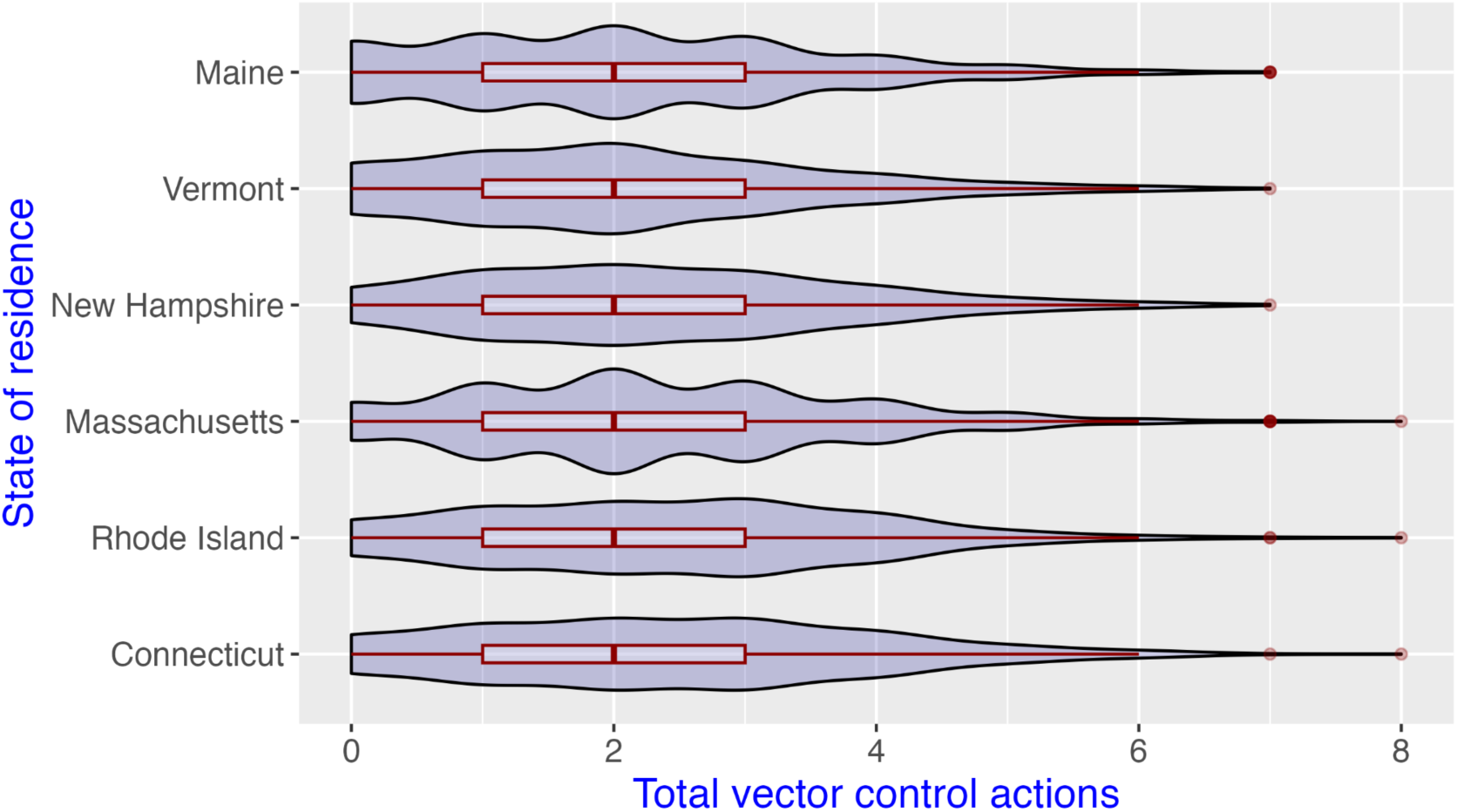
Distribution of total self-reported vector control actions, by state, “Is Tick Control Helping” (ITCH) survey respondents, New England, USA, 2023-2024 (N = 4,957).

**Table S1A.** Prevalence of specific self-reported vector control actions by state, “Is Tick Control Helping” (ITCH) survey respondents, New England, USA, 2023 survey questions (N = 3,926).

|  | Survey total<br>responding<br>“yes” | State-level total responses |  |  |  |  |  |
| --- | --- | --- | --- | --- | --- | --- | --- |
|  |  | ME<br>N=1,482 | NH<br>N=313 | VT<br>N=452 | RI<br>N=361 | MA<br>N=1,140 | CT<br>N=178 |
|  | N | (% of state-level total responding “yes”) |  |  |  |  |  |
| **2023 vector control: Rodent bait box** | 416 | (10.6) | (8.3) | (8.2) | (15.8) | (10.0) | (14.0) |
| **2023 vector control: Tick Tubes** | 521 | (13.4) | (14.4) | (13.7) | (9.1) | (14.0) | (12.4) |
| **2023 vector control: Vegetation removal** | 1,501 | (39.3) | (38.7) | (41.4) | (34.3) | (36.1) | (42.1) |
| **2023 vector control: Tick-safe landscaping (woodchips, moving play structures, etc.)** | 730 | (17.5) | (19.5) | (14.8) | (18.8) | (19.8) | (27.0) |
| **2023 vector control: Removal of items that store water around the property** | 1,408 | (53.7) | (70.3) | (61.9) | (69.3) | (74.3) | (70.2) |
| **2023 vector control: Deer fencing** | 328 | (8.8) | (6.7) | (7.1) | (9.1) | (7.3) | (15.7) |
| **2023 vector control: Light candles, torches, or fires as repellents** | 1,191 | (27.5) | (32.6) | (31.6) | (30.7) | (31.8) | (37.1) |
| **2023 vector control: Apply pesticides on my property myself (or a member of my household)** | 814 | (19.8) | (20.1) | (19.9) | (20.8) | (21.5) | (26.4) |
| **2023 vector control: Hire a commercial business to apply pesticides** | 569 | (12.3) | (13.1) | (2.7) | (31.3) | (16.2) | (20.2) |
| **2023 vector control: None** | 487 | (16.9) | (9.9) | (15.0) | (9.1) | (8.2) | (6.2) |

**Table S1B.** Prevalence of specific self-reported vector control actions by state, “Is Tick Control Helping” (ITCH) survey respondents, New England, USA, 2024 survey questions (N = 929)

|  | Survey total responding “yes”<br>N | State-level total responses |  |  |  |  |  |
| --- | --- | --- | --- | --- | --- | --- | --- |
|  |  | ME<br>N=10 | NH<br>N=29 | VT<br>N=8 | RI<br>N=133 | MA<br>N=439 | CT<br>N=310 |
|  |  | (% of state-level total responding “yes”) |  |  |  |  |  |
| **2024 tick control: Rodent bait box** | 121 | (10.0) | (13.8) | (12.5) | (20.3) | (10.5) | (13.5) |
| **2024 tick control: Tick Tubes** | 100 | (10.0) | (6.9) | (12.5) | (9.8) | (12.3) | (9.4) |
| **2024 tick control: Vegetation removal** | 253 | (40.0) | (44.8) | (37.5) | (28.6) | (28.2) | (22.9) |
| **2024 tick control: Tick-safe landscaping (woodchips, moving play structures, etc)** | 143 | (20.0) | (20.7) | (0) | (12.8) | (15.7) | (15.8) |
| **2024 tick control: Deer fencing** | 69 | (0) | (13.8) | (0) | (12.0) | (5.5) | (8.1) |
| **2024 tick control: Apply pesticides on my property myself (or a member of my household)** | 90 | (20.0) | (13.8) | (12.5) | (12.0) | (8.7) | (9.4) |
| **2024 tick control: Hire a commercial business to apply pesticides** | 280 | (20.0) | (27.6) | (0) | (39.1) | (27.1) | (31.9) |
| **2024 tick control: Leaf removal** | 421 | (30.0) | (48.3) | (50.0) | (50.4) | (44.2) | (44.8) |
| **2024 tick control: Other** | 58 | (20.0) | (6.9) | (0) | (6.0) | (6.6) | (5.5) |
| **2024 mosquito control: Removal of items that store water around the property** | 642 | (70.0) | (69.0) | (37.5) | (69.2) | (74.5) | (62.3) |
| **2024 mosquito control: Light candles, torches, or fires as repellents** | 345 | (40.0) | (34.5) | (12.5) | (30.8) | (33.9) | (45.2) |
| **2024 mosquito control: Apply pesticides on my property myself (or a member of my household)** | 106 | (30.0) | (10.3) | (12.5) | (12.0) | (10.5) | (11.9) |
| **2024 mosquito control: Hire a commercial business to apply pesticides** | 141 | (10.0) | (17.2) | (0) | (24.8) | (14.4) | (12.6) |
| **2024 mosquito control: Other** | 78 | (10.0) | (6.9) | (0) | (5.3) | (10.9) | (6.5) |

**Table S2A.** Prevalence of self-reported personal prevention actions, “Is Tick Control Helping” (ITCH) survey respondents, New England, USA, 2023 survey questions (N = 3,926). Highest prevalence categories in boldface.

|  | Overall<br>N | State |  |  |  |  |  |
| --- | --- | --- | --- | --- | --- | --- | --- |
|  |  | ME<br>N=1,482 | NH<br>N=313 | VT<br>N=452 | RI<br>N=361 | MA<br>N=1,140 | CT<br>N=178 |
|  |  | (%) | (%) | (%) | (%) | (%) | (%) |
| **Wear long pants/long sleeves outdoors** |  |  |  |  |  |  |  |
| Always | 1033 | (26.8) | (26.5) | (25.9) | (19.9) | (27.5) | (28.7) |
| Sometimes | **2791** | **(71.5)** | **(70.9)** | **(71.0)** | **(77.0)** | **(69.6)** | **(65.7)** |
| Never | 101 | (1.8) | (2.6) | (3.1) | (3.0) | (2.8) | (5.6) |
| NA | 1 | (0) | (0) | (0) | (0) | (0.1) | (0) |
| **Apply insect repellent to clothing or wear commercially-treated clothing (permethrin)** |  |  |  |  |  |  |  |
| Always | 930 | (24.2) | (27.2) | (25.2) | (16.6) | (24.5) | (19.1) |
| Sometimes | **2284** | **(60.5)** | **(55.6)** | **(59.1)** | **(56.8)** | **(56.1)** | **(57.3)** |
| Never | 688 | (14.7) | (16.9) | (15.3) | (26.0) | (18.7) | (23.0) |
| NA | 24 | (0.7) | (0.3) | (0.4) | (0.6) | (0.7) | (0.6) |
| **Apply insect repellent to skin (DEET)** |  |  |  |  |  |  |  |
| Always | 612 | (15.8) | (14.4) | (17.0) | (13.9) | (15.8) | (14.6) |
| Sometimes | **2655** | **(67.0)** | **(72.8)** | **(62.4)** | **(69.3)** | **(68.2)** | **(69.7)** |
| Never | 646 | (16.9) | (12.8) | (20.1) | (16.9) | (15.7) | (14.0) |
| NA | 13 | (0.3) | (0) | (0.4) | (0) | (0.3) | (1.7) |
| **Shower after spending time outdoors** |  |  |  |  |  |  |  |
| Always | 1365 | (32.9) | (36.7) | (36.1) | (35.2) | (35.8) | (36.5) |
| Sometimes | **2342** | **(61.3)** | **(57.8)** | **(59.1)** | **(61.5)** | **(57.5)** | **(60.1)** |
| Never | 213 | (5.7) | (5.1) | (4.6) | (3.3) | (6.4) | (3.4) |
| NA | 6 | (0.1) | (0.3) | (0.2) | . | (0.3) | . |
| **Do tick checks** |  |  |  |  |  |  |  |
| Always | **2789** | **(75.0)** | **(84.7)** | **(71.0)** | **(59.3)** | **(66.7)** | **(65.7)** |
| Sometimes | 1102 | (24.6) | (14.7) | (28.1) | (39.3) | (32.1) | (32.0) |
| Never | 34 | (0.3) | (0.6) | (0.9) | (1.4) | (1.2) | (2.2) |
| NA | 1 | (0.1) | (0) | (0) | (0) | (0) | (0) |
| **Treat pets with pesticide products** |  |  |  |  |  |  |  |
| Always | **1767** | **(48.5)** | **(43.5)** | **(53.5)** | **(42.4)** | **(39.0)** | **(40.4)** |
| Sometimes | 479 | (11.7) | (12.8) | (13.9) | (12.2) | (12.5) | (9.6) |
| Never | 436 | (10.1) | (15.0) | (7.5) | (13.0) | (12.5) | (9.6) |
| NA | 1244 | (29.8) | (28.8) | (25.0) | (32.4) | (36.1) | (40.4) |
| **Tuck in cuffs and socks** |  |  |  |  |  |  |  |
| Always | 745 | (20.8) | (18.2) | (21.7) | (13.3) | (17.9) | (16.9) |
| Sometimes | **2112** | **(55.1)** | **(56.5)** | **(55.1)** | **(52.6)** | **(51.8)** | **(49.4)** |
| Never | 1058 | (24.0) | (25.2) | (23.0) | (33.8) | (29.7) | (33.1) |
| NA | 11 | (0.1) | (0) | (0.2) | (0.3) | (0.5) | (0.6) |

**Table S2B.** Prevalence of self-reported personal prevention actions, “Is Tick Control Helping” (ITCH) survey respondents, New England, USA, 2024 survey questions (N = 929). Highest prevalence categories in boldface.

|  | Overall<br>N | State |  |  |  |  |  |
| --- | --- | --- | --- | --- | --- | --- | --- |
|  |  | ME<br>N=10 | NH<br>N=29 | VT<br>N=8 | RI<br>N=133 | MA<br>N=439 | CT<br>N=310 |
|  |  | (%) | (%) | (%) | (%) | (%) | (%) |
| **Wear long pants/long sleeves outdoors** |  |  |  |  |  |  |  |
| Always | 204 | **(60.0)** | (24.1) | (25.0) | (16.5) | (23.9) | (20.0) |
| Sometimes | **674** | (10.0) | **(72.4)** | **(75.0)** | **(75.2)** | **(72.4)** | **(73.5)** |
| Never | 50 | (30.0) | (3.4) | (0) | (8.3) | (3.4) | (6.5) |
| NA | 1 | (0) | (0) | (0) | (0) | (0.2) | (0) |
| **Apply insect repellent to clothing or wear commercially-treated clothing (permethrin)** |  |  |  |  |  |  |  |
| Always | 177 | (20.0) | (27.6) | (0) | (21.1) | (21.2) | (14.8) |
| Sometimes | **525** | **(60.0)** | **(58.6)** | **(62.5)** | **(54.1)** | **(54.4)** | **(60.0)** |
| Never | 219 | (20.0) | (13.8) | (37.5) | (24.1) | (23.2) | (24.5) |
| NA | 8 | (0) | (0) | (0) | (0.8) | (1.1) | (0.6) |
| **Apply insect repellent to skin (DEET)** |  |  |  |  |  |  |  |
| Always | 145 | (10.0) | (20.7) | (12.5) | (15.8) | (15.7) | (15.2) |
| Sometimes | **626** | **(80.0)** | **(69.0)** | **(37.5)** | **(67.7)** | **(67.4)** | **(67.4)** |
| Never | 154 | (10.0) | (10.3) | **(50.0)** | (15.8) | (16.4) | (17.1) |
| NA | 4 | (0) | (0) | (0) | (0.8) | (0.5) | (0.3) |
| **Shower after spending time outdoors** |  |  |  |  |  |  |  |
| Always | 413 | (20.0) | (37.9) | **(50.0)** | **(50.4)** | (40.1) | **(49.4)** |
| Sometimes | **471** | **(70.0)** | **(58.6)** | **(50.0)** | (44.4) | **(54.9)** | (46.1) |
| Never | 45 | (10.0) | (3.4) | (0) | (5.3) | (5.0) | (4.5) |
| **Do tick checks** |  |  |  |  |  |  |  |
| Always | **590** | **(60.0)** | **(72.4)** | **(100.0)** | **(59.4)** | **(66.1)** | **(60.0)** |
| Sometimes | 315 | (40.0) | (27.6) | (0) | (36.8) | (31.4) | (37.4) |
| Never | 23 | (0) | (0) | (0) | (3.8) | (2.5) | (2.3) |
| NA | 1 | (0) | (0) | (0) | (0) | (0) | (0.3) |
| **Treat pets with pesticide products** |  |  |  |  |  |  |  |
| Always | **342** | **(20.0)** | (27.6) | **(37.5)** | **(39.8)** | **(35.5)** | **(38.7)** |
| Sometimes | 130 | **(20.0)** | (10.3) | **(37.5)** | (12.0) | (12.8) | (16.1) |
| Never | 145 | (10.0) | **(34.5)** | (12.5) | (9.8) | (14.1) | (18.7) |
| NA | 312 | (50.0) | (27.6) | (12.5) | (38.3) | (37.6) | (26.5) |
| **Tuck in cuffs and socks** |  |  |  |  |  |  |  |
| Always | 138 | (20.0) | (24.1) | (0) | (11.3) | (16.2) | (13.9) |
| Sometimes | **476** | **(40.0)** | **(55.2)** | **(62.5)** | **(57.1)** | **(51.3)** | **(48.4)** |
| Never | 311 | (40.0) | (20.7) | (37.5) | (30.8) | (32.3) | (37.1) |
| NA | 4 | (0) | (0) | (0) | (0.8) | (0.2) | (0.6) |

**Table S3.**
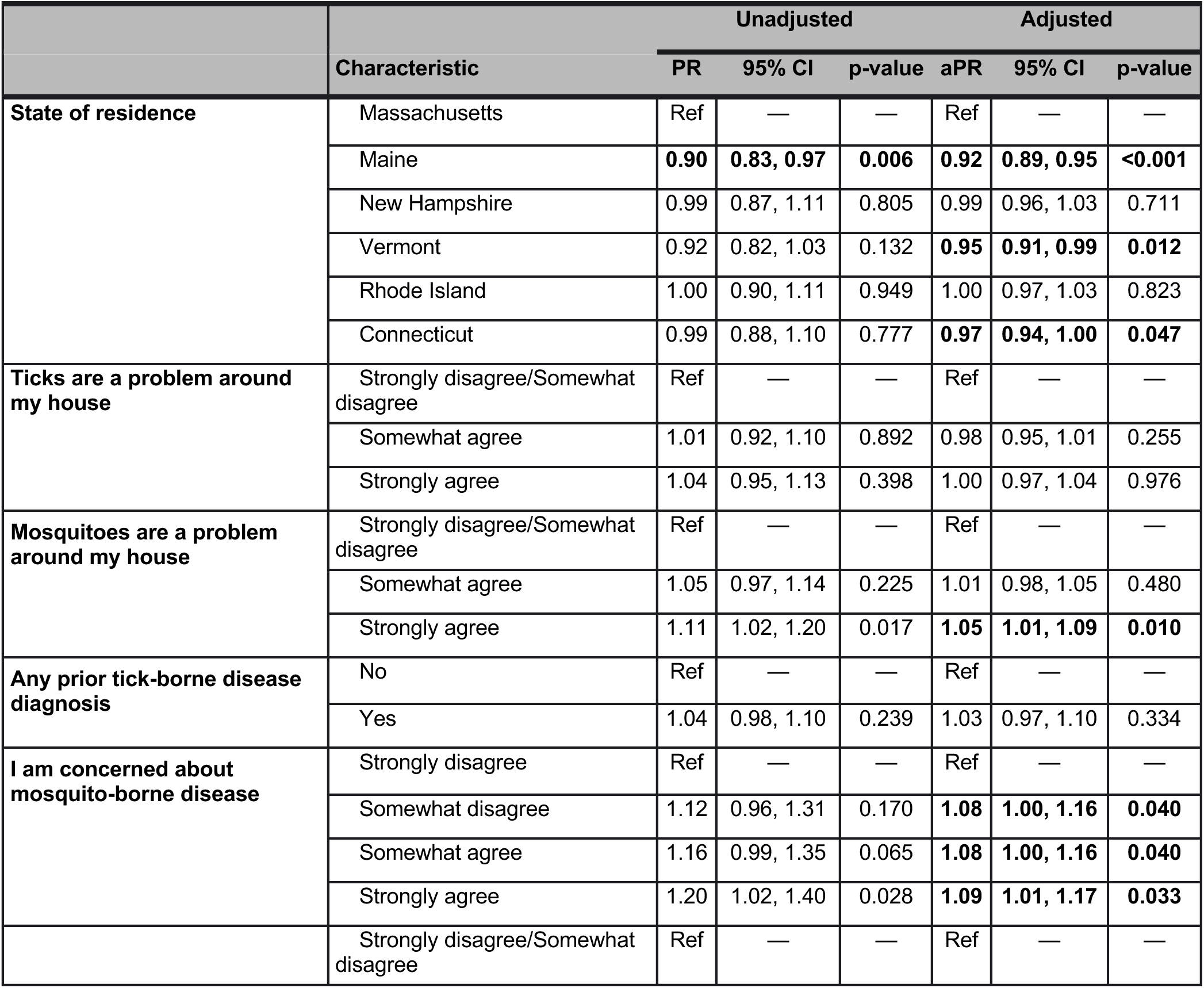
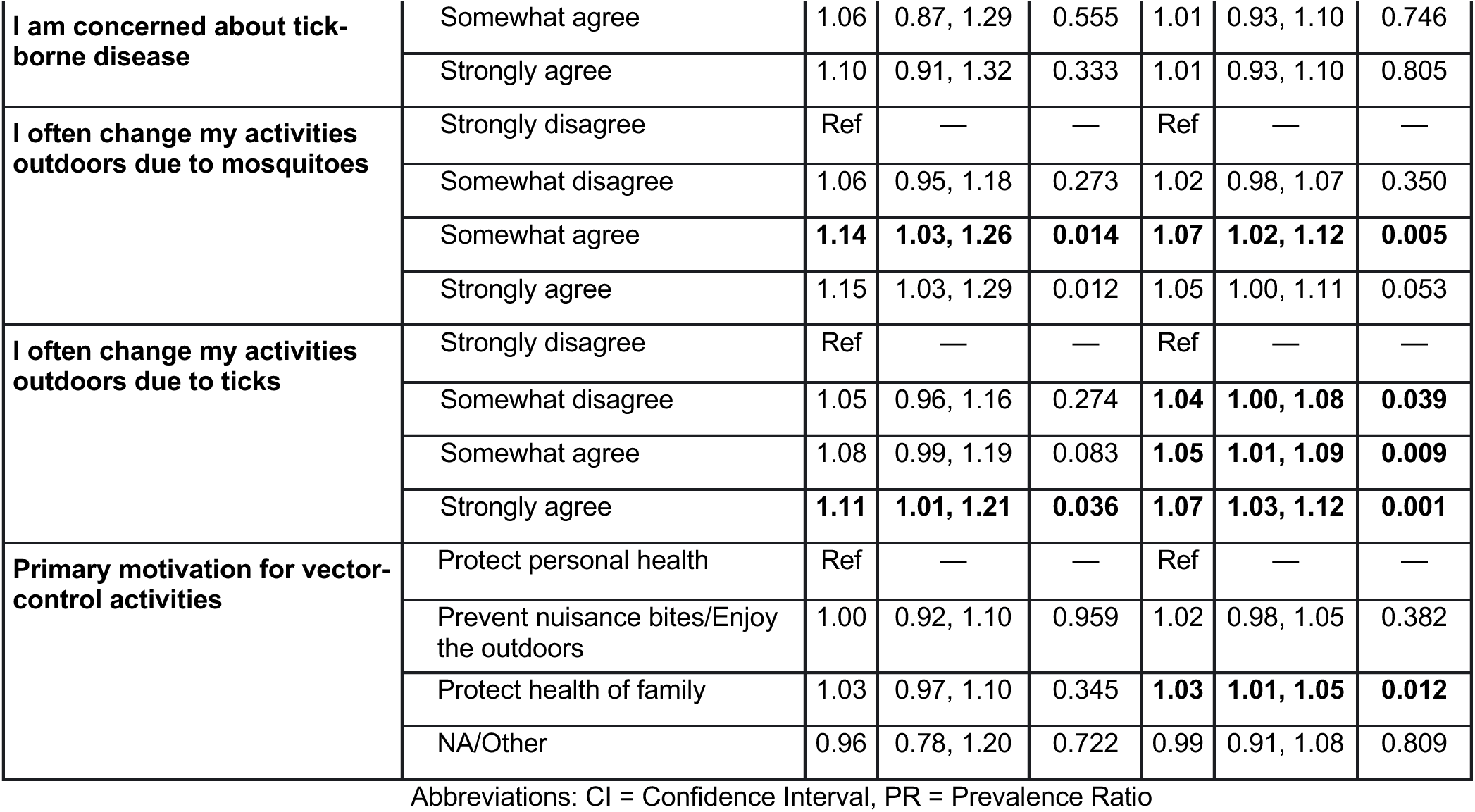
Multivariable robust Poisson models to estimate prevalence ratios (PRs), for use of any self-reported vector control intervention at the household level, New England, 2023-2024. Note: final model adjusted for age, survey year, sex/gender, yard size, property type, and household income. Values in boldface are statistically significant with p < 0.05.

|  | Characteristic | Unadjusted |  |  | Adjusted |  |  |
| --- | --- | --- | --- | --- | --- | --- | --- |
|  |  | PR | 95% CI | p-value | aPR | 95% CI | p-value |
| **State of residence** | Massachusetts | Ref | — | — | Ref | — | — |
|  | Maine | **0.90** | **0.83, 0.97** | **0.006** | **0.92** | **0.89, 0.95** | **<0.001** |
|  | New Hampshire | 0.99 | 0.87, 1.11 | 0.805 | 0.99 | 0.96, 1.03 | 0.711 |
|  | Vermont | 0.92 | 0.82, 1.03 | 0.132 | **0.95** | **0.91, 0.99** | **0.012** |
|  | Rhode Island | 1.00 | 0.90, 1.11 | 0.949 | 1.00 | 0.97, 1.03 | 0.823 |
|  | Connecticut | 0.99 | 0.88, 1.10 | 0.777 | **0.97** | **0.94, 1.00** | **0.047** |
| **Ticks are a problem around my house** | Strongly disagree/Somewhat disagree | Ref | — | — | Ref | — | — |
|  | Somewhat agree | 1.01 | 0.92, 1.10 | 0.892 | 0.98 | 0.95, 1.01 | 0.255 |
|  | Strongly agree | 1.04 | 0.95, 1.13 | 0.398 | 1.00 | 0.97, 1.04 | 0.976 |
| **Mosquitoes are a problem around my house** | Strongly disagree/Somewhat disagree | Ref | — | — | Ref | — | — |
|  | Somewhat agree | 1.05 | 0.97, 1.14 | 0.225 | 1.01 | 0.98, 1.05 | 0.480 |
|  | Strongly agree | 1.11 | 1.02, 1.20 | 0.017 | **1.05** | **1.01, 1.09** | **0.010** |
| **Any prior tick-borne disease diagnosis** | No | Ref | — | — | Ref | — | — |
|  | Yes | 1.04 | 0.98, 1.10 | 0.239 | 1.03 | 0.97, 1.10 | 0.334 |
| **I am concerned about mosquito-borne disease** | Strongly disagree | Ref | — | — | Ref | — | — |
|  | Somewhat disagree | 1.12 | 0.96, 1.31 | 0.170 | **1.08** | **1.00, 1.16** | **0.040** |
|  | Somewhat agree | 1.16 | 0.99, 1.35 | 0.065 | **1.08** | **1.00, 1.16** | **0.040** |
|  | Strongly agree | 1.20 | 1.02, 1.40 | 0.028 | **1.09** | **1.01, 1.17** | **0.033** |
|  | Strongly disagree/Somewhat disagree | Ref | — | — | Ref | — | — |
| I am concerned about tick-borne disease | Somewhat agree | 1.06 | 0.87, 1.29 | 0.555 | 1.01 | 0.93, 1.10 | 0.746 |
|  | Strongly agree | 1.10 | 0.91, 1.32 | 0.333 | 1.01 | 0.93, 1.10 | 0.805 |
| I often change my activities outdoors due to mosquitoes | Strongly disagree | Ref | — | — | Ref | — | — |
|  | Somewhat disagree | 1.06 | 0.95, 1.18 | 0.273 | 1.02 | 0.98, 1.07 | 0.350 |
|  | Somewhat agree | **1.14** | **1.03, 1.26** | **0.014** | **1.07** | **1.02, 1.12** | **0.005** |
|  | Strongly agree | 1.15 | 1.03, 1.29 | 0.012 | 1.05 | 1.00, 1.11 | 0.053 |
| I often change my activities outdoors due to ticks | Strongly disagree | Ref | — | — | Ref | — | — |
|  | Somewhat disagree | 1.05 | 0.96, 1.16 | 0.274 | **1.04** | **1.00, 1.08** | **0.039** |
|  | Somewhat agree | 1.08 | 0.99, 1.19 | 0.083 | **1.05** | **1.01, 1.09** | **0.009** |
|  | Strongly agree | **1.11** | **1.01, 1.21** | **0.036** | **1.07** | **1.03, 1.12** | **0.001** |
| Primary motivation for vector-control activities | Protect personal health | Ref | — | — | Ref | — | — |
|  | Prevent nuisance bites/Enjoy the outdoors | 1.00 | 0.92, 1.10 | 0.959 | 1.02 | 0.98, 1.05 | 0.382 |
|  | Protect health of family | 1.03 | 0.97, 1.10 | 0.345 | **1.03** | **1.01, 1.05** | **0.012** |
|  | NA/Other | 0.96 | 0.78, 1.20 | 0.722 | 0.99 | 0.91, 1.08 | 0.809 |

Abbreviations: CI = Confidence Interval, PR = Prevalence Ratio

**Table S4.** Multivariable Poisson models for total self-reported vector control interventions at the household level, New England, 2023-2024, comparing northern and southern regions within New England. Notes: IRR = incidence rate ratio; final model adjusted for respondant age, sex/gender, survey year, yard size, property type, and household income. State was combined into a binary of Northern and Southern states in New England. Values in boldface are statistically significant with p < 0.05.

| Variable | Level | Unadjusted |  |  | Adjusted |  |  |
| --- | --- | --- | --- | --- | --- | --- | --- |
|  |  | IRR | 95% CI | p-value | aIRR | 95% CI | p-value |
| Residence in Northern or Southern New England | Southern New England | Ref | — | — | Ref | — | — |
|  | Northern New England | **0.89** | **0.85, 0.92** | **<0.001** | **0.90** | **0.86, 0.94** | **<0.001** |
| Ticks are a problem around my house | Strongly disagree/Somewhat disagree | Ref | — | — | Ref | — | — |
|  | Somewhat agree | **1.12** | **1.06, 1.19** | **<0.001** | 1.04 | 0.98, 1.11 | 0.180 |
|  | Strongly agree | **1.25** | **1.18, 1.32** | **<0.001** | **1.09** | **1.02, 1.17** | **0.011** |
| Mosquitoes are a problem around my house | Strongly disagree/Somewhat disagree | Ref | — | — | Ref | — | — |
|  | Somewhat agree | **1.19** | **1.12, 1.25** | **<0.001** | **1.08** | **1.02, 1.14** | **0.013** |
|  | Strongly agree | **1.36** | **1.29, 1.44** | **<0.001** | **1.15** | **1.07, 1.23** | **<0.001** |
| Any prior tick-borne disease diagnosis | No | Ref | — | — | Ref | — | — |
|  | Yes | **1.09** | **1.05, 1.13** | **<0.001** | **1.07** | **1.03, 1.12** | **<0.001** |
| I am concerned about tick-borne disease | Strongly disagree/Somewhat disagree | Ref | — | — | Ref | — | — |
|  | Somewhat agree | **1.16** | **1.51, 1.93** | **0.029** | 1.02 | 0.89, 1.17 | 0.744 |
|  | Strongly agree | **1.33** | **1.17, 1.51** | **<0.001** | 1.00 | 0.87, 1.15 | 0.987 |
| **I am concerned about mosquito-borne disease** | Strongly disagree | Ref | — | — | Ref | — | — |
|  | Somewhat disagree | **1.19** | **1.07, 1.32** | **0.002** | 1.10 | 0.99, 1.23 | 0.089 |
|  | Somewhat agree | **1.33** | **1.20, 1.48** | **<0.001** | **1.15** | **1.03, 1.29** | **0.014** |
|  | Strongly agree | **1.52** | **1.37, 1.70** | **<0.001** | **1.24** | **1.10, 1.39** | **<0.001** |
| **I often change my activities outdoors due to mosquitoes** | Strongly disagree | Ref | — | — | Ref | — | — |
|  | Somewhat disagree | **1.14** | **1.06, 1.23** | **<0.001** | 1.04 | 0.97, 1.13 | 0.277 |
|  | Somewhat agree | **1.27** | **1.19, 1.36** | **<0.001** | 1.08 | 0.99, 1.17 | 0.070 |
|  | Strongly agree | **1.46** | **1.36, 1.57** | **<0.001** | **1.12** | **1.03, 1.23** | **0.012** |
| **I often change my activities outdoors due to ticks** | Strongly disagree | Ref | — | — | Ref | — | — |
|  | Somewhat disagree | **1.09** | **1.03, 1.17** | **0.005** | 1.05 | 0.98, 1.13 | 0.135 |
|  | Somewhat agree | **1.26** | **1.19, 1.34** | **<0.001** | **1.16** | **1.11, 1.28** | **<0.001** |
|  | Strongly agree | **1.34** | **1.27, 1.43** | **<0.001** | **1.20** | **1.11, 1.29** | **<0.001** |
| **Primary motivation for vector-control activities** | Protect personal health | Ref | — | — | Ref | — | — |
|  | Prevent nuisance bites/ Enjoy the outdoors | 1.01 | 0.95, 1.07 | 0.829 | 1.05 | 0.98, 1.11 | 0.156 |
|  | Protect health of family | **1.15** | **1.10, 1.20** | **<0.001** | **1.12** | **1.07, 1.17** | **<0.001** |
|  | NA/Other | 0.90 | 0.78, 1.04 | 0.155 | 0.95 | 0.82, 1.10 | 0.497 |

Abbreviations: CI = Confidence Interval, IRR = Incidence Rate Ratio

## Notes

### Competing Interest Statement

The authors have declared no competing interest.

### Author Declarations

The study protocol was reviewed and approved by the University of Massachusetts Amherst Institutional Review Board (IRB) as the central IRB (Protocol # 3969).

